# Repeated population-based surveys of antibodies against SARS-CoV-2 in Southern Brazil

**DOI:** 10.1101/2020.05.01.20087205

**Authors:** Mariângela F Silveira, Aluísio J D Barros, Bernardo L Horta, Lúcia C Pellanda, Odir A Dellagostin, Claudio J Struchiner, Marcelo N Burattini, Andréia R M Valim, Evelise M Berlezi, Jeovany M Mesa, Maria Letícia R Ikeda, Marilia A Mesenburg, Marina Mantesso, Marinel M Dall’Agnol, Raqueli A Bittencourt, Fernando P Hartwig, Ana M B Menezes, Fernando C Barros, Pedro C Hallal, Cesar G Victora

**Affiliations:** Universidade Federal de Pelotas, Brazil; Fundação Universidade Federal de Ciências de Saúde de Porto Alegre, Brazil; Fundação Getúlio Vargas and Universidade do Estado do Rio de Janeiro, Brazil; Universidade de São Paulo, Brazil and Universidade Federal de São Paulo, Brazil; Universidade de Santa Cruz do Sul, Brazil; Universidade de Ijuí, Brazil; IMED - Passo Fundo, Brazil; Universidade do Vale do Rio dos Sinos, Brazil; Universidade de Caxias do Sul, Brazil; Universidade Federal de Santa Maria, Brazil; Secretaria Municipal de Saúde de Uruguaiana, Brazil

**Author notes:** Joint last authors (Pedro C Hallal, Cesar G Victora). **Correspondence to:** Pedro C Hallal, Marechal Deodoro 1160 – 96020-220 - Pelotas, RS, Brazil –.

## Abstract

Population based data on COVID-19 are urgently needed for informing policy decisions, yet few such studies are available anywhere, as most surveys rely on self-selected volunteers. In the Brazilian State of Rio Grande do Sul (population 11.3 million), we are carrying out fortnightly household surveys in nine of the largest cities. Multi-stage probability sampling was used in each city to select 500 households, within which one resident was randomly chosen for testing. The Wondfo lateral flow rapid test for detecting antibodies against SARS-CoV-2 has been validated in four different settings, including our own, with pooled estimates of sensitivity (84.8%, 95% CI 81.4%;87.8%) and specificity (99.0%, 95% CI 97.8%;99.7%), which are within the acceptable range for epidemiological studies. In the first wave of the study (April 11-13), 4,188 subjects were tested, of whom two were positive (0.0477%; 95% confidence interval (CI) 0.0058%;0.1724%). In the second round (Apr 25-27) there were six positive subjects (0.1333%; 95% CI 0.0489%;0.2900%). We also tested family members of positive index cases, and nine out of 19 had positive results. Testing of reported COVID-19 cases according to RT-PCR confirmed that the test was highly sensitive under field conditions. The epidemic is at an early stage in the State, as the first case was reported on Feb 28, and by Apr 30, 50 deaths were registered. Strict lockdown measures were implemented in mid-March, and our results suggest that compliance was high, with full or near full compliance rates of 79.4% in the first and 71.7% in the second round. As far as we know, this is the only large population anywhere undergoing regular household serological surveys for COVID-19. The results show that the epidemic is at an early phase, and findings from the next rounds will allow us to document time trends and propose Public Health measures.

## INTRODUCTION

Despite calls for population-based data on COVID-19,^1^ there have been remarkably few household seroprevalence surveys anywhere, and none in Latin America.^2^ In Rio Grande do Sul, the southernmost state in Brazil (population 11.3 million), the first case of COVID-19 was diagnosed on February 29, 2020. As of April 30,1,466 confirmed cases (129 per 1,000,000 inhabitants) and 50 deaths had been reported (http://ti.saude.rs.gov.br/covidl9/). It is important to remark that in the state, as also in Brazil, only persons with moderate to severe symptoms had been tested, using PCR to detect SARS-Cov-2. The state and most municipal governments issued strong social distancing policies in mid-March, including closures of schools, shops and services, except for businesses deemed to be essential.

Other than studies based on convenience samples, such as individuals who volunteered to be tested, supermarket customers, or blood donors, there are few general population sample surveys in the literature. In a national study in Iceland^3^, one of the three groups of participants was recruited through random sampling of the population, but only about one third of those invited were tested. In this group, 13 of 2283 persons tested positive (0.6%; 95% CI 0.3;0.9) in quantitative real-time polymerase-chain-reaction (qRT-PCR) assays. A national household study in Austria used random sampling to invite households to participate. Of the households contacted, 77% or 2,197 declared their willingness to participate, and 1,541 persons were successfully tested. Assuming two persons per household, those tested correspond to about 30% of the intended sample. The study found a prevalence of 0.33% (95% CI 0.12;0.76%.) using qRT-PCR, or 5 individuals out of those tested.^4^. Smaller studies in hot spots for COVID-19 showed prevalence of 14% the German city of Gangelt^5^ and 3% in the Italian village of Vò.^6^

As expected, studies based on volunteers found higher prevalence, as was the case for the first study in Iceland (0.9%)^3^, the population screening in South Korea (2.1%)^7^ and two studies in California: prevalence of 1.5% in Santa Clara county^8^ and 4.1% in Los Angeles County. http://publichealth.lacountv.gov/phcommon/public/media/mediapubhpdetail.cfm?prid=2328

Starting on April 11-13, we set out to test the presence of antibodies against SARS-CoV-2 in population-based samples of 500 individuals in each of nine sentinel cities in the state, with a total sample of 4,500. The same methodology was used in a second round in the same cities on April 25-27, and subsequent rounds are planned to take place every two weeks in order to monitor how the pandemic is evolving.

## METHODS

The state of Rio Grande do Sul is divided by the National Institute of Geography and Statistics in eight intermediary regions (Figure 1). The main city in each region was selected for the study. In the main metropolitan region, we selected the State Capital, Porto Alegre, and Canoas, the second largest city in the metropolitan area. Populations ranged from 78,915 in Ijuí to 1,409,351 in Porto Alegre (https://cidades.ibge.gov.br/brasil/rs/panorama).

**Figure 1.**
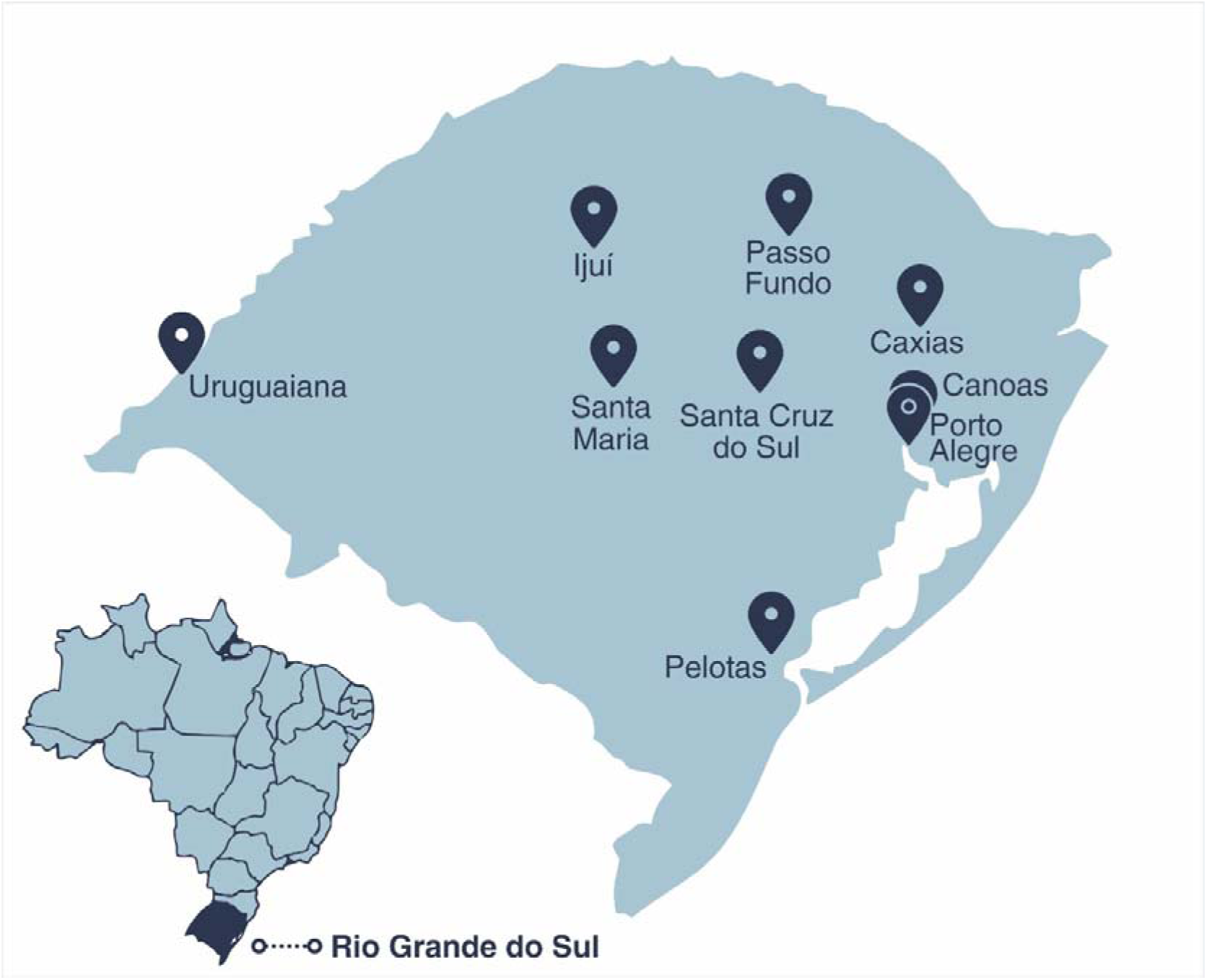
Location of the nine sentinel cities.

We used multistage sampling to select 50 census tracts with probability proportionate to size in each sentinel city, and 10 households at random in each tract based on census listings updated in 2019. All household members were listed at the beginning of the visit, and one individual was randomly selected through an app used for data collection. The survey waves took place on April 10-12 and 25-27.

The Statewide sample of 4,500 individuals allows estimating a prevalence levels of 3% and 10% with margins of error of 0.5 and 1.0 percent points, respectively.

In the first wave, interviewers had listings of 35 households in each tract. Any refusals at household level led to selection of the next household in the list, and so on until 10 households were included. In the second wave, field workers went to the house visited in the first wave, and then selected the tenth household to its right. In case of refusal, the next household to the right side was selected. In the case of acceptance at the household level but the index individual refused to provide a sample, a second member was selected. If this person also refused, the field workers moved on to the next household in the list.

Prevalence of antibodies was assessed with a rapid test using finger prick blood samples - the WONDFO SARS-CoV-2 Antibody Test (Wondfo Biotech Co., Guangzhou, China). This test detects immunoglobulins of both IgG and IgM isotypes specific to SARS-CoV-2 antigens in a lateral flow assay. The capture reagent consists of an unspecified viral antigen immobilized at a defined position on a nitrocellulose membrane. Following the introduction of the sample, a solution containing labelled detector anti-immunoglobulin monoclonal antibodies is added. If the test is valid, a control line appears on the kit’s window. If this line is not visible, the test is deemed inconclusive, which is very uncommon. A positive result is triggered by binding of the detector antibody to any serum immunoglobulins immobilized on the viral antigen, and is visible as a second colored line. Two drops of blood from a pinprick are sufficient to detect the presence of antibody.

Four independent validation studies are available for the rapid test. Its sensitivity and specificity are 86.4% and 99.6% according to the manufacturer, using samples collected from 361 confirmed cases and 235 negative controls(https://en.wondfo.com.cn/product/wondfo-sars-cov-2-antibodv-test-lateral-flow-method-2/). The tests were purchased in bulk by the Brazilian government, being earmarked for use in population surveys and surveillance programs. An initial validation study was carried out by the National Institute for Quality Control in Health (INCQS, Oswaldo Cruz Foundation, RJ, Brazil) using 18 qRT-PCR positive and 77 negative serum samples. The reported sensitivity was 100.0% (95% confidence interval (Cl) 81.5;100.0%), while specificity was 98.7% (95% Cl 93.0;100.9%). Recently, Whitman and colleagues^9^ evaluated 10 different lateral flow assays using as specimens plasma or serum samples from symptomatic SARSCoV-2 RT-PCR-positive individuals and 108 pre-COVID-19 negative controls. Sensitivity of the Wondfo test was 81.5% (95% Cl 70.0-90.1%) among 65 patients with a positive RT-PCR 11 or more days before the test, and specificity was 99.1% (95% Cl 94.9;100.0%). Of the 10 tests studied, the Wondfo test was one of the two lateral flow tests with the best performance. Lastly, we carried out our own validation study, based on 83 volunteers with a positive qRT-PCR result 10 days or more before the rapid test. This analysis showed a sensitivity of 77.1% (95% Cl 66.6;85.6%). We also analysed 100 sera samples collected in 2012 from participants of the 1982 Pelotas (Brazil) Birth Cohort Study^10^ and found 98 negative results, yielding a specificity estimate of 98.0% (95% Cl 93.0;99.8%). By pooling the results from the four separate validations studies, weighted by sample sizes, sensitivity is estimated at 84.8% (95% Cl 81.4%;87.8%) and specificity at 99.0% (95% Cl 97.8%;99.7%).

Participants answered short questionnaires including sociodemographic information (sex, age, schooling and skin color), COVID-19-related symptoms, use of health services, compliance with social distancing measures and use of masks. Field workers used tablets or smartphones to record the full interviews, register all answers, and photograph the test results. All positive or inconclusive tests were read by a second observer, as well as 20% of the negative tests. If the index subject in a household had a positive result, all other family members were invited to be tested.

Interviewers were tested and found to be negative for the virus, and were provided with individual protection equipment that was discarded after visiting each home. Ethical approval was obtained from the Brazilian’s National Ethics Committee (process number 30415520.2.0000.5313), with written informed consent from all participants. Positive cases were reported to the statewide COVID-19 surveillance system. The study protocol was published prior to the first wave of data collection.^11^ Data will become publicly available upon request from the corresponding author 30 days after publication.

In the analyses reported in the body of this article, we analyzed the surveys as if it they included simple random samples from the population, using the exact binomial method for confidence intervals. We calculated absolute (in percent points) and relative differences between the two survey waves regarding the prevalence of infection. P-values were calculated using Cochran’s Q. heterogeneity test, implemented as fixed-effects meta-regression, which also yielded confidence intervals for the differences. More complex analyses with allowances for the sampling design, population weights and corrections for the specificity and sensitivity of the rapid test, are included in the web annex. All analyses were performed using R version 3.6.1 (https://www.r-project.org/). The “metafor” package was used to compare the prevalence between surveys.

## RESULTS

Out of the planned 4,500 interviews, it was possible to test 4,188 individuals in the first round. The number of tests carried out included 500 in each of five cities (Pelotas, Passo Fundo, Santa Cruz, Caxias), 396 in Porto Alegre, 332 in Canoas, 499 in Uruguaiana and 461 in Santa Maria. In the last three cities, the desired sample size was not completed due to logistic difficulties resulting from the need to complete the survey in a 3-day period. Refusals, requiring the selection of the next household in the census tract listing, ranged from 5.4% in ljui to 26.9% in Santa Maria, with a median of 17.9% in the nine cities. In the second round, it was possible to obtain 500 interviews in each of the nine cities.

Table 1 shows the characteristics of individuals who provided blood samples. Both samples were similar in terms of sex, age, skin color and schooling distributions. Although the nine sentinel cities are not representative of the state as a whole, the comparison shows what the samples had higher proportions of women and of older persons than the state as a whole. Young children were particularly underrepresented. Up-to-date information on schooling is not available for the State.

**Table 1.**
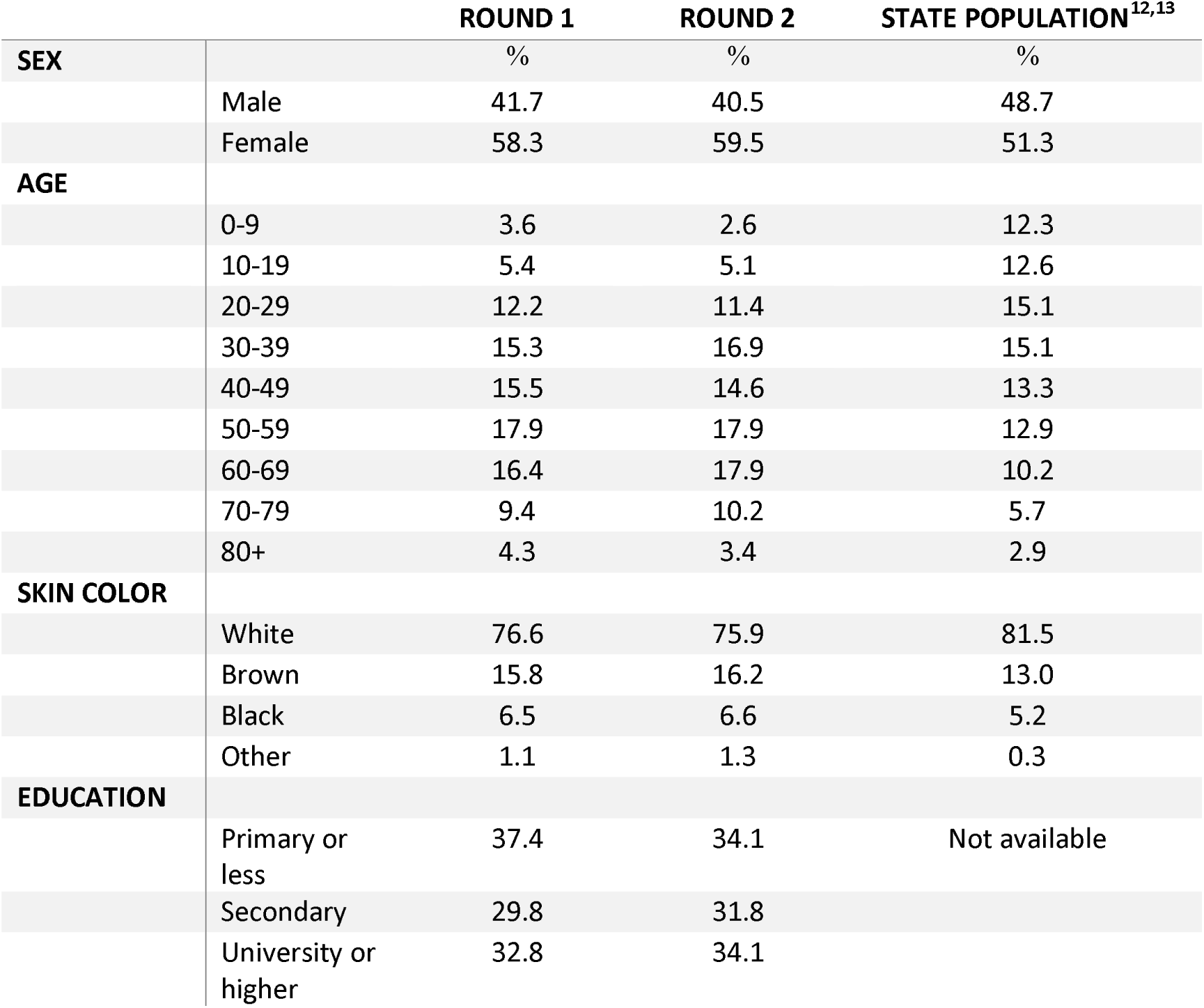
Sociodemographic characteristics of the two samples in nine cities and of the State population.

Of the 4,188 individuals tested in the first round, 10 had inconclusive results and only two (0.0477%; 95% confidence interval (Cl) 0.0058%;0.1724%) tested positive, one each in the cities of Pelotas and Uruguaiana. In the second round, there were two inconclusive results and six positive subjects (0.1333%; 95% Cl 0.0489%;0.2900%). The absolute prevalence difference between the second and first waves was equal 0.086 percent point (95% Cl −0.400;0.211; P=0.181), and the ratio was equal to 2.793 (95% Cl 0.564;13.831, P=0.208).

Given the small numbers of subjects who tested positive, we focus the presentation on unadjusted results. The web annex provides results from more complex analyses, all of which produced results that are highly comparable to those reported here.

Regarding social distancing measures, 20.6% of respondents reported leaving home on a daily basis, 58.3% leaving home occasionally for essential activities, and 21.1% staying at home all the time in the first phase, and 28.3%, 53.4% and 18.3%, respectively, in the second phase.

The households of the eight positive cases in the two phases included other 20 residents. Of these, 19 were tested; the rapid test showed nine positive, eight negative, and two inconclusive results. One positive case lived alone. Among the other seven, four had at least another positive individual in their families.

As an additional check on how the rapid test performed under field work conditions, we conducted two separate assessments. The first was during the validation study in Porto Alegre, where 83 RT-PCR positive individuals were tested in the field using the rapid test. As described in the Methods section, 64 of these had positive results with the rapid test. Second, a more limited assessment entailed asking the coordinators of field work in different cities whether they were aware of any RT-PCR positive individuals in their communities. Four persons were identified and tested, all of whom had positive results in the rapid test.

## DISCUSSION

This is the first report on repeated population-based surveys for the detection of SARS-CoV-2 antibodies. With two weeks interval we were able to perform antibody tests on representative samples in nine sentinel cities in Rio Grande do Sul State in Southern Brazil.

Based on reported death rates by April 30, 2020, Rio Grande do Sul is one of the six states, out of 27, with the lowest mortality, of 4 per million, well below Rio de Janeiro (46 per million) or Sao Paulo (49 per million). Amazonas state (92 per million) shows the highest death rates. The national mortality rate is estimated at 26 per million. (https://covid.saude.gov.br/)

Taking our present results at face value, there would be 477 cases per million inhabitants (95% confidence interval 58-1,719 cases) in the first wave, compared to 62 reported cases per million, as of April 14. According to the results of the second wave (April 25-27), there would be 1,333 cases per million inhabitants (95% Cl 489;2900) compared to 128 reported cases per million as of April 30. Additional estimates, taking into account corrections for the sample design, population weighting and adjustment for sensitivity and specificity, are provided in the web annex.

Important concerns have been issued about rapid serological tests, but these mostly refer to their use in making clinical decisions,^14^ and on issuing “immunity passports” ^15^ for individuals who are assumed to have developed immunity. Both of these circumstances refer to individual-level diagnoses based on rapid tests. Use of rapid tests for population-based estimates, and particularly for monitoring trends over time, is a different issue for which rapid tests with less than perfect sensitivity and specificity may be acceptable. The Wondfo lateral flow test used in our analyses underwent four different validation studies, being able to correctly identify 5 out of every 6 RT-PCR confirmed cases, and 99 out of 100 individuals without SARS-CoV-2 antibodies. Among 10 lateral flow tests recently assessed by Whitman and colleagues^9^, it was among the two with the best performance. Our finding of positive results for 10 of 13 family members of the six index individuals who tested positive confirms that the performance of the rapid test was adequate under field conditions.

The limitations of our analyses include the restriction of the sample to sentinel cities that jointly account for 31% of the state’s population, while smaller towns and rural areas were not included. Second, antibody tests result in many false negatives for recent infections, particularly within the first two weeks since contagion, and thus prevalence reflects levels of infection a week or two prior to the survey, about 15 days after the first case was reported in the state. The non-response rate at household level, estimated at 17.9%, was low compared to other population-based studies,^3,4^, or to studies based on volunteers. Our samples had fewer children than expected, which was probably due to their reluctance to undergo a finger prick when randomly selected within the household; in these cases, a second person was randomly selected and if that person also refused the household was replaced.

Lastly, our results were at the lower range of the 95% confidence interval for the false positive rate, which in the pooled estimate from four validation studies was estimated at 1.0% (95% Cl 0.3%;2.2%). In these studies, specificity was measured in frozen samples. Whitman and colleagues, in their analyses of 10 lateral flow tests, observed “*moderate-to-strong positive bands in several pre-COVlD-19 blood donor specimens, some of them positive by multiple assays, suggesting the possibility of non-specific binding of plasma proteins, non-specific antibodies, or cross-reactivity with other viruses.”*^9^ Our results on family clustering show that - out of the seven index cases who lived alone - four had family members who also tested positive. These four individuals are most likely true positives, thus suggesting that up to four of the remaining index cases with positive results, out of 8,689 individuals, might represent false positive results. The test’s specificity would then be equal to 99.95% (95% Cl 99.88;99.99%).

Our finding of low prevalence is consistent with an early phase of the pandemic, coupled with high compliance with social distancing measures, as confirmed by our own results. Such a low prevalence level is compatible with other population-based studies: 0.6% in Iceland ^3^ and 0.3% in Austria,^4^ which is close to Northern Italy which was strongly hit by the pandemic. One should note that in both studies about 2/3 of those invited failed to participate, compared to our own non-response rate of 17.9%. Our results are not comparable with those based on self-selected volunteers.

The surveys are being partly funded by the state and national governments of Brazil. Survey results were disseminated, two days after the completion of data collection round, in press briefings with the presence of the state governor, who is making use of the information to guide stay-at-home and other policies. Results from the next rounds of our study - planned for May 8-10 and 22-23 - will allow us to follow the dynamics of the pandemic in the state, especially when social restriction measures are starting to be relaxed in most municipalities.

## Data Availability

The full dataset will be made publicly available no longer than 30 days after each wave of data collection.

## Notes

**Funding:** This work started through the Data Committee created by the State of Rio Grande do Sul government to fight the COVID-19 pandemics. The tests used in the study have been provided by the Brazilian Ministry of Health. Funding for data collection was provided by UNIMED Porto Alegre, Instituto Cultural Floresta and Instituto Serrapilheira.

### Competing Interest Statement

The authors have declared no competing interest.

### Clinical Protocols

http://www.cienciaesaudecoletiva.com.br/artigos/evolucao-da-prevalencia-de-infeccao-por-covid19-no-rio-grande-do-sul-inqueritos-sorologicos-seriados/17547

### Funding Statement

This work started through the Data Committee created by the State of Rio Grande do Sul government to fight the COVID-19 pandemics. The tests used in the study have been provided by the Brazilian Ministry of Health. Funding for data collection was provided by UNIMED Porto Alegre, Instituto Cultural Floresta and Instituto Serrapilheira.

## References

1. Pearce N, Vandenbroucke JP, VanderWeele TJ, Greenland S. Accurate Statistics on COVID-19 Are Essential for Policy Guidance and Decisions. American journal of public health 2020: el-e3.

2. Barreto ML, Barros AJD, Carvalho MS, et al. [What is urgent and necessary to inform policies to deal with the COVID-19 pandemic in Brazil?]. Rev Bras Epidemiol 2020; 23: e200032.

3. Gudbjartsson DF, Helgason A, Jonsson H, et al. Spread of SARS-CoV-2 in the Icelandic Population. The New England journal of medicine 2020.

4. Institute for Social Research and Consulting Ogris & Hofinger GmbH (SORA). COVID-19 Prevalence, 2020.

5. Regalado A. Blood tests show 14% of people are now immune to covid-19 in one town in Germany. 2020. https://www.technologvreview.com/2020/04/09/999015/blood-tests-show-15-of-people-are-now-immune-to-covid-19-in-one-town-in-germanv/ (accessed April 27, 2020.

6. Day M. Covid-19: identifying and isolating asymptomatic people helped eliminate virus in Italian village. BMJ 2020; 368: m1165.

7. Ministry of Health and Welfare (South Korea). Coronavirus disease 19, Repubic of South Korea. 2020. http://ncov.mohw.go.kr/en.

8. Bendavid E, Mulaney B, Sood N, et al. COVID-19 Antibody Seroprevalence in Santa Clara County, California. medRxiv 2020: 2020.04.14.20062463.

9. Whitman JD, Hiatt J, Mowery CT, et al. Test performance evaluation of SARS-CoV-2 serological assays, *(unpublished)* 2020.

10. Horta BL, Gigante DP, Goncalves H, et al. Cohort profile update: the 1982 Pelotas (Brazil) Birth Cohort Study. International journal of epidemiology 2015; 44(2): 441-e.

11. Hallal P, Horta B, Barros A, et al. Evolução da prevalencia de infecção por COVID-19 no Rio Grande do Sul: inquèritos sorológicos seriados. Cien Saude Colet 2020.

12. Institute Brasileiro de Geografia e Estatística. Pesquisa nacional por amostra de domicNios: síntese de indicadores 2015. Rio de Janeiro: IBGE, 2016.

13. Institute Brasileiro de Geografia e Estatfstica. Projeções da População. 2019. HTTPS://WWW.IBGE.GOV.BR/ESTATISTICAS/SOCIAIS/POPULACAO/9109-PROJECAO-DA-POPULACAO.HTML?=&T=RESULTADOS (accessed April 30, 2020.

14. World Health Organization. Advice on the use of point-of-care immunodiagnostic tests for COVID-19. 2020. https://www.who.int/news-room/commentaries/detail/advice-on-the-use-of-point-of-care-immunodiagnostic-tests-for-covid-19 (accessed April 30, 2020.

15. World Health Organization. “Immunity passports” in the context of COVID-19. 2020. https://www.who.int/news-room/commentaries/detail/immunity-passports-in-the-context-of-covid-19 (accessed April 30, 2020.

